# Examining the Burden of Non-Communicable Diseases among Elderly Individuals Aged 70 and Older in Jordan over Three Decades

**DOI:** 10.1101/2024.06.11.24308789

**Authors:** Aya Abdelhaq, Duaa Barakat, Renad Zghoul, Iyad Sultan

## Abstract

**Background:** This study investigates the trends in non-communicable diseases (NCDs) among the elderly population aged 70 and above in Jordan from 1991 to 2022. The elderly Jordanian population was targeted as it has vastly increased over the years, being approximately 6 times higher in 2021 than in 1991. The aim of this study is to provide a comprehensive insight on the burden of NCDs among Jordan’s elderly population .

**Methods:** Using data extracted from the Global Burden of Disease (GBD), incidence rates, disability-adjusted life years (DALYs), and mortality rates associated with NCDs were analyzed.

**Results:** The findings of this study reveal a significant increase in the total burden of NCDs over the three decades reaching an absolute DALYs number of above 262,000 by 2021. Elderly females appear to have a slightly higher burden of NCDs when compared to males over the specified time period of 1991 to 2021. Cardiovascular diseases and diabetes were identified as the primary contributors to this increase and seem to have the highest two death rate ranks. However, when compared to global rates Jordan appears to have a 34% lower NCDs burden. The incidence rates appear to be stable with minimal change suggesting that the NCDs incidence has remained relatively constant in Jordan over the past three decades.

**Conclusion:** This upward trend underscores the escalating public health challenge posed by NCDs in Jordan’s aging population. This study provides a comprehensive overview of the changes in NCD burden and aims to inform healthcare policy and strategic planning to mitigate the impact of NCDs on the elderly in Jordan.

## Introduction

Non-communicable diseases (NCDs) refer to chronic diseases mainly including cardiovascular diseases, diabetes, cancer, and chronic respiratory diseases. The World Health Organization (WHO) states that NCDs are considered to be the leading cause of death worldwide as they tend to kill 41 million people each year (74% of all deaths globally) (1). NCDs are usually caused by a combination of environmental (air pollution), behavioral (tobacco use, physical inactivity, unhealthy diet), and metabolic (high blood pressure, obesity, hyperglycemia) risk factors (1).

Based on the Jordan National Stepwise Survey (STEPS) for Non-communicable Disease (NCD) Risk Factors (2019) (2), NCDs were found to be the leading cause of morbidity and mortality in Jordan, accounting for 78% of deaths. The data also reported the prevalence of hypertension (52%), diabetes (20%) and high risk for cardiovascular disease (25%) among adults aged 45 to 69 years old (2). Additionally, Jordan is classified by the World Bank as a lower middle-income country (3). According to WHO more than three quarters of global NCD deaths occur in low- and middle-income countries (1).

Aging has also been identified as a risk factor to the development of NCDs as the elderly population have a higher prevalence of NCDs (4). This is due to the fact that the elderly tend to have multiple comorbidities, longer duration of chronic diseases, and accumulated exposure to risk factors (5). All countries have been facing a significant rapid increase in the aging population, increasing the prevalence of NCDs worldwide (6). Accordingly, the aging population in Jordan has been reported to be increasing (7). As indicated in Figure 1., a substantial increase was shown from the years of 1991 to 2021 among the total elderly population in Jordan, including all age groups from 70 years and above, with the combined population increasing by approximately 6 times from 1991 to 2021 (see Appendix). Identifying the trends of NCDs among the elderly may potentially aid in reducing its burden on the Jordanian population. This allows public health systems in Jordan to implement strategies that help them combat the diseases of high burden, providing the appropriate public health needs for the population.

**Figure 1.**
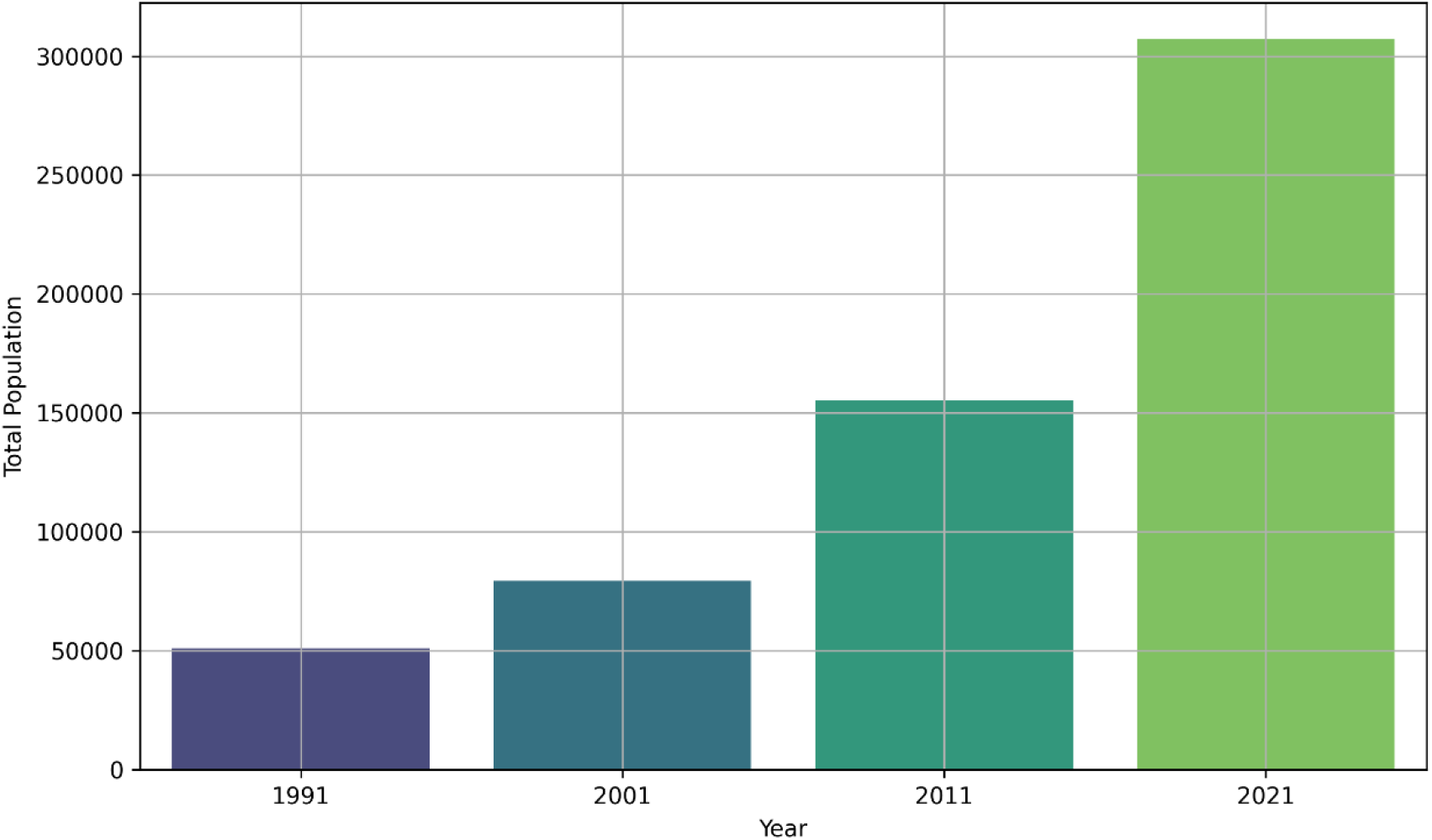
Elderly population trends in Jordan (1991-2021)

Being the largest and most comprehensive global research program of disease burden, the Global Burden of Disease (GBD) quantifies levels and trends in health that can be referred to for improving health systems and comparing each country’s progress along with identifying the main causes of health loss that can be prevented (8). It provides data on the estimates of mortality, deaths, years of life lost due to premature mortality (YLLs), years lived with disability (YLDs), and disability-adjusted life years (DALYs) due to major diseases in addition to the various risk factors and injuries. The data is currently available for the years 1990 to 2021 and includes a total of 204 countries and territories (9).

The objective of this study is to identify the trends in the burden of non-communicable diseases between the years of 1991 and 2021 in Jordan. The study focuses on the elderly population aged 70 years and above. The current data from the GBD was analyzed including the NCDs incidence rates, DALYs, and death rates. The analysis was performed over the years of 1991 to 2021 due to the availability of GBD data up until 2021, allowing the examination of the trend changes over an extended timeframe (9). To our information, no previous study was conducted in Jordan to study the burden of NCDs for this specific age group, over the years of 1991 to 2021, using the GBD data. Additionally, this study uniquely analyzes data from the years 2020 and 2021 which were only recently added by the GBD. This study provides a comprehensive insight on the burden of NCDs in Jordan’s elderly population. Accordingly, the study aimed to answer the following research questions:

> **RQ1**. How have the DALYs numbers changed over time for NCDs?
>
> **RQ2**. How do the DALYs for NCDs in Jordan compare to the global rates across all ages?
>
> **RQ3**. How have the trends of NCDs among the elderly population changed over time in terms of incidence rates?
>
> **RQ4**. How have the causes of death rates among the elderly population changed over time, in regard to NCDs?

To this end, the following sections in this study include a review of the currently available literature, the methodology section describes the approach followed to obtain the data and address the research questions, and the findings are presented in the results section. The findings are then discussed in detail, finally concluding with a summary of the primary findings and future research recommendations.

## Literature Review

Non-communicable diseases (NCDs) are a major global health issue, accounting for 60% of deaths. As cardiovascular and chronic obstructive diseases increase, they pose significant risks to health and productivity. Epidemiologists play a crucial role in implementing methods to identify preventable causes, monitor disease occurrence, and ensure the effectiveness of prevention programs (10). Gong et al. (2018) studied the global burden of NCDs and their impact on developing countries (11). NCDs, including cardiovascular diseases, cancer, diabetes, obesity, and chronic respiratory diseases, account for a sizable portion of global deaths and disease burden, especially in developing countries. By 2020, NCDs are predicted to account for 70% of the global burden of disease and seven out of every ten deaths in developing countries. Preventive measures and public health initiatives are crucial for controlling the spread of NCDs. Additionally, the Arab world has observed an increase in the incidence of non-communicable illnesses, including obesity and other risk factors. Despite this, policy responses have been lacking, with variances among nations and poor implementation (12). Among the elderly population, NCDs are considered a significant health concern, with a mortality rate reported in 2012 to be 533/10 million. These diseases account for 86.6% of total deaths, with cardiovascular diseases, cancer, and chronic respiratory diseases being the main causes (11). A systematic analysis was conducted for the GBD 2019 study estimating mortality and disability trends for the population aged 70 years and above and evaluating patterns in causes of death, disability, and risk factors (13). The study involved elderly participants from 204 countries and territories from 1990 to 2019. The primary outcome measures included years of life lost, years lived with disability, disability-adjusted life years, life expectancy at age 70 (LE-70), healthy life expectancy at age 70 (HALE-70), the proportion of years in ill health at age 70 (PYIH-70), risk factors, and data coverage index. The results showed that globally, the population of older adults has increased since 1990, and all-cause death rates have decreased for both genders. However, mortality rates due to falls increased between 1990 and 2019, mainly because of reductions in non-communicable diseases. Disability burden was primarily driven by functional decline, vision and hearing loss, and symptoms of pain. Life expectancy at the age of 70 has continued to rise globally, mostly due to decreases in chronic diseases. Adults aged 70 years and above living in high-income countries and regions with better healthcare access and quality were found to experience the highest and most healthy life expectancies. Until now, no study has previously addressed the burden of NCDs among individuals aged 70 years and above specifically in Jordan from 1991 to 2021. Therefore, to address this gap, this research aims to study the burden of NCDs among the Jordanian elderly population, in addition to comparing it to the global countries, due to the observed increase in the elderly population in Jordan.

## Methods

### Data Source

The Institute for Health Metrics and Evaluation (IHME) has organized the global burden of disease (GBD) which evaluates the burden of 371 diseases, including estimates of incidence, prevalence, mortality, years of life lost (YLLs), years lived with disability (YLDs), and disability-adjusted life-years (DALYs), and healthy life expectancy (HALE) from 1991 to 2021 across 204 countries and territories for both sexes (14). Level 2 NCDs featured in GBD 2021 include cardiovascular diseases, neoplasms (cancers), chronic respiratory diseases, diabetes, neurological disorders, digestive diseases, mental disorders, musculoskeletal disorders, and other non-communicable diseases (including sense organ diseases, skin and subcutaneous diseases, and substance use disorders) (14). Measures’ counts and age-standardized rates are calculated globally for seven super-regions, 21 regions, 204 countries and territories (including 21 countries with subnational locations), and 811 subnational locations from 1990 to 2021 (14).

In this research, the required input data are extracted from the most recent edition of GBD 2021 (10). The data included the population size, NCD incidence rates, death rates, and DALYs, for the years of 1991 to 2021, specifically for the Jordanian population aged 70 years and above.

### DALYs

The GBD calculates YLDs through multiplying the cause-age-sex-location-year-specific prevalence of sequelae by their respective disability weights for each disease and injury. YLLs are calculated by multiplying cause-age-sex-location-year-specific deaths by the standard life expectancy at the age that death occurred. DALYs are calculated by summing YLDs and YLLs.

### Incidence rate

Incidence rate is a crucial epidemiological metric used in Global Burden of Disease (GBD) studies to quantify the number of new cases of a certain disease within a specified population and period providing a more comprehensive understanding of the spread of disease. It is calculated by dividing the number of new cases during a specific period by the population at risk during the same period and multiplying by 10^n, where ‘n’ indicates the scale at which the incidence rate is expressed, usually per 1000, 10,000, or 100,000 person-years (14).

### Death rate

The Death Rate is calculated by dividing the number of deaths within a specific population during a given period by the total population during the same time period. It reflects the number of deaths relative to the size of the population. It can indicate various factors such as the prevalence of diseases, the effectiveness of healthcare systems, living conditions, and overall quality of life (14).

### Word bank rating

The World Bank categorizes countries into income groups based on Gross National Income (GNI) per capita, which is used in GBD studies to assess health outcomes and disease burdens. This classification helps in understanding how economic development influences health outcomes and disease burdens. The income groups are defined as follows: low-income, lower-middle, upper-middle, and high-income economies, with GNI per capita ranging from $1,045 to $12,696. Jordan is a country that is considered to be in the lower-middle-income group. In this research, Jordan’s NCD burden was compared to the other income categories (14).

## Results

The following section presents the results of the analysis conducted on the data obtained from the GBD, offering comprehensive insights on the trends and burden of NCDs among the elderly population in Jordan to answer the proposed research questions.

### NCDs Burden over time

Figure 2. presents a line graph showing a consistent upward trend in the total DALYs number for NCDs in Jordan over the years of 1991 to 2021. The graph begins at below 100,000 and constantly increases, eventually exceeding a total number of over 250,000 DALYs by 2021. The growth rate appears to be significantly accelerating. This suggests that NCDs have been causing a greater impact on the elderly population over time.

**Figure 2.**
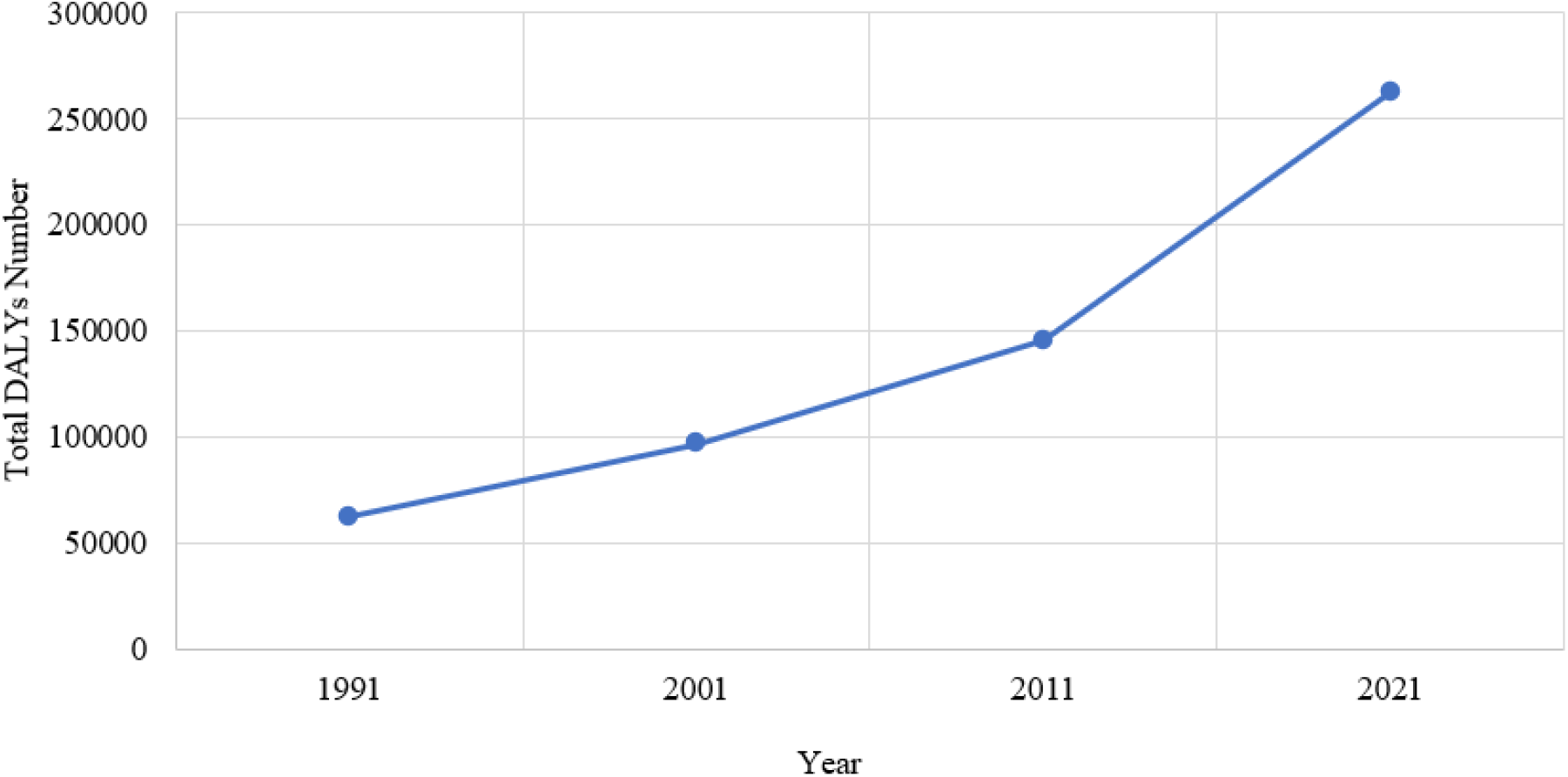
Total DALYs numbers for elderly NCDs in Jordan over time.

Figure 3. presents the changes in DALYs numbers across the 4 time points for both genders of the Jordanian population across all ages. There appears to be a significant increase among both genders with only a slightly higher increase in males indicating a more rapid growth in the burden of NCDs compared to females. This shows a significant rise in NCDs from 1991 to 2021, indicating a growing public health issue requiring specific measures to overcome it.

**Figure 3.**
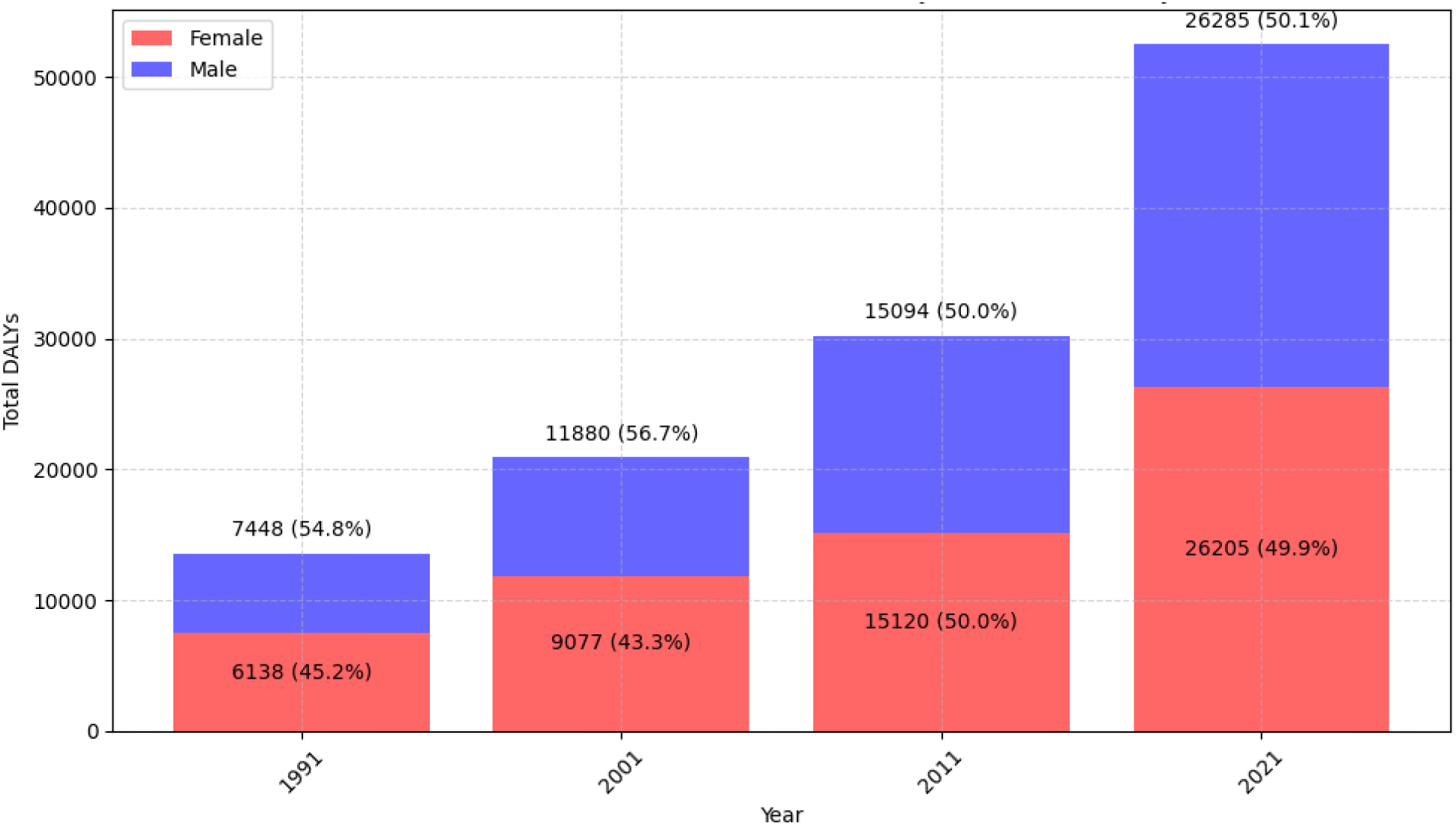
Change in DALYs numbers for NCDs in Jordan over time (by sex).

Table 1. displays the percentage change in the DALYs rate for the different NCDs among both genders of the elderly population over the specific time period of 1991 to 2021. The data significantly points towards a reduction in the prevalence or cases of many diseases over the period covered. Cardiovascular diseases appear to have the highest contribution to the overall NCDs. Some disease categories like diabetes and kidney diseases and musculoskeletal disorders show slight increases over the years, indicating areas that might need focused health interventions. The elderly females seem to have a higher burden of NCDs than males throughout 1991 to 2021.

**Table 1.** Change in DALYs rate per 100,000 individuals aged 70 years and above in Jordan over time (by cause and sex).

| Cause | Sex | 1991 | 2001 | 2011 | 2021 | Changes In Rate % |
| --- | --- | --- | --- | --- | --- | --- |
| Non-communicable diseases | Both | 62840.50 | 96883.40 | 145284.77 | 262555.37 | -0.32 |
| Non-communicable diseases | Female | 36041.82 | 56604.83 | 72990.07 | 135003.82 | -0.34 |
| Non-communicable diseases | Male | 26798.68 | 40278.57 | 72294.70 | 127551.55 | -0.27 |
| Cardiovascular diseases | Both | 32054.26 | 47785.53 | 60616.11 | 102642.98 | -0.48 |
| Cardiovascular diseases | Female | 19023.29 | 29530.96 | 31475.49 | 53884.32 | -0.51 |
| Cardiovascular diseases | Male | 13030.97 | 18254.57 | 29140.61 | 48758.66 | -0.43 |
| Chronic respiratory diseases | Both | 2997.25 | 4459.85 | 5955.67 | 10295.49 | -0.44 |
| Chronic respiratory diseases | Female | 1334.50 | 2094.72 | 2378.56 | 4339.68 | -0.43 |
| Chronic respiratory diseases | Male | 1662.75 | 2365.14 | 3577.12 | 5955.82 | -0.46 |
| Diabetes and kidney diseases | Both | 8311.06 | 15159.82 | 25186.87 | 48288.10 | -0.03 |
| Diabetes and kidney diseases | Female | 5340.94 | 9766.08 | 13499.47 | 25736.54 | -0.14 |
| Diabetes and kidney diseases | Male | 2970.12 | 5393.73 | 11687.40 | 22551.56 | 0.18 |
| Digestive diseases | Both | 2813.38 | 4162.34 | 5014.53 | 8067.41 | -0.53 |
| Digestive diseases | Female | 1646.42 | 2483.22 | 2637.29 | 4245.71 | -0.55 |
| Digestive diseases | Male | 1166.96 | 1679.12 | 2377.24 | 3821.70 | -0.50 |
| Mental disorders | Both | 954.71 | 1465.15 | 2763.00 | 5565.76 | -0.03 |
| Mental disorders | Female | 585.65 | 844.06 | 1538.29 | 3236.71 | -0.03 |
| Mental disorders | Male | 369.06 | 621.09 | 1224.71 | 2329.06 | -0.01 |
| Musculoskeletal disorders | Both | 2648.54 | 4182.60 | 8348.18 | 16838.01 | 0.06 |
| Musculoskeletal disorders | Female | 1597.83 | 2365.20 | 4596.28 | 9608.32 | 0.06 |
| Musculoskeletal disorders | Male | 1050.71 | 1817.40 | 3751.90 | 7229.70 | 0.08 |
| Neoplasms | Both | 4807.25 | 7729.77 | 15522.54 | 26871.84 | -0.08 |
| Neoplasms | Female | 2082.94 | 3528.20 | 6316.00 | 11538.93 | -0.02 |
| Neoplasms | Male | 2724.31 | 4201.58 | 9206.54 | 15332.91 | -0.14 |
| Neurological disorders | Both | 3748.15 | 5259.94 | 9531.92 | 19678.27 | -0.15 |
| Neurological disorders | Female | 2021.92 | 2667.34 | 4629.67 | 10225.83 | -0.13 |
| Neurological disorders | Male | 1726.23 | 2592.60 | 4902.25 | 9452.44 | -0.16 |
| Other non-communicable diseases | Both | 1254.17 | 1964.29 | 3671.26 | 7139.35 | -0.05 |
| Other non-communicable diseases | Female | 670.47 | 981.84 | 1753.05 | 3506.22 | -0.08 |
| Other non-communicable diseases | Male | 583.70 | 982.45 | 1918.21 | 3633.13 | -0.02 |
| Sense organ diseases | Both | 2920.90 | 4190.39 | 7670.72 | 15179.76 | -0.15 |
| Sense organ diseases | Female | 1544.43 | 2048.80 | 3643.63 | 7606.85 | -0.15 |
| Sense organ diseases | Male | 1376.48 | 2141.59 | 4027.09 | 7572.91 | -0.15 |
| Skin and subcutaneous diseases | Both | 299.27 | 473.80 | 911.39 | 1820.97 | 0.01 |
| Skin and subcutaneous diseases | Female | 178.97 | 272.77 | 484.24 | 1003.24 | -0.02 |
| Skin and subcutaneous diseases | Male | 120.30 | 201.02 | 427.14 | 817.73 | 0.06 |
| Substance use disorders | Both | 31.55 | 49.91 | 92.58 | 167.42 | -0.13 |
| Substance use disorders | Female | 14.45 | 21.63 | 38.10 | 71.49 | -0.14 |
| Substance use disorders | Male | 17.10 | 28.28 | 54.47 | 95.93 | -0.13 |

### DALYs Comparison: NCDs in Jordan vs. Global Rates

Table 2. compares the burden of different NCDs in Jordan to global and World Bank income groups in 2021, calculating percentage differences of Jordan in relation to these groups across all ages. Both higher and lower burdens of NCDs were examined in Jordan compared to global and income group averages. This may aid in identifying the areas where targeted interventions seem to be effective and those where interventions are needed to address the health challenges and reduce the burden of those diseases in Jordan. There seems to be variations among different NCDs with a particularly higher burden of diabetes and kidney diseases in Jordan in comparison to global averages, low-income, and lower-middle-income groups. Mental disorders appear to have a higher burden in Jordan when compared to global averages and all income categories except for the high-income group. Musculoskeletal disorders also seem to have a higher burden in Jordan when compared to low- and lower-middle-income categories. In comparison to both global and low-income country averages, substance use disorders appear to have a significantly lower burden in Jordan. Cardiovascular diseases appear to be significant contributors to the overall NCD burden in Jordan however, they have a lower burden in comparison to the global averages and the income group classifications. Also, the burden of chronic respiratory diseases, digestive diseases, skin and subcutaneous diseases, and neoplasms is lower in Jordan compared to global and income group averages. These findings indicate the importance of strengthening healthcare systems and implementing tailored health interventions in aim to manage and control NCDs of high burden in Jordan. Implementing preventive measures focusing on lifestyle changes, allocating resources efficiently to maintain low burdens in areas where Jordan is performing better, and developing and enforcing health policies that address the specific needs of the elderly population may potentially aid in managing the health impacts of Jordan’s aging population and reducing the burden of NCDs.

**Table 2.** Comparison between Jordan and DALYs rates per 100,000 individuals of Global and Word Bank income categories across all ages in 2021 (both sexes).

| Cause | Global | Jordan | High Income | Low Income | Lower Middle Income | Upper Middle Income | Jordan vs Global % Difference | Jordan vs Low-Income % Difference | Jordan vs Lower Middle Income % Difference | Jordan vs Upper Middle Income % Difference | Jordan vs High Income % Difference |
| --- | --- | --- | --- | --- | --- | --- | --- | --- | --- | --- | --- |
| Non-communicable diseases | 65658.89 | 43113.17 | 80464.3 | 47292.32 | 60173.8 | 70731.8 | -34.34 | -8.84 | -28.35 | -39.05 | -46.42 |
| Cardiovascular diseases | 16278.32 | 7374.02 | 15241.86 | 9576.86 | 16160.93 | 18755.35 | -54.7 | -23 | -54.37 | -60.68 | -51.62 |
| Chronic respiratory diseases | 4123.99 | 1160.32 | 4105.99 | 2300.41 | 4593.59 | 4038.18 | -71.86 | -49.56 | -74.74 | -71.27 | -71.74 |
| Diabetes and kidney diseases | 4702.44 | 4819.9 | 5633.16 | 3206.76 | 4572.04 | 4838.27 | 2.5 | 50.3 | 5.42 | -0.38 | -14.44 |
| Digestive diseases | 3420.53 | 1225.78 | 3457.33 | 3216.66 | 3807.64 | 2972.65 | -64.16 | -61.89 | -67.81 | -58.76 | -64.55 |
| Mental disorders | 5909.76 | 6943.14 | 7350.28 | 5463.85 | 5646.93 | 5681.67 | 17.49 | 27.07 | 22.95 | 22.2 | -5.54 |
| Musculoskeletal disorders | 6157.23 | 5448.77 | 9704.11 | 3289.97 | 5081.26 | 6637.89 | -11.51 | 65.62 | 7.23 | -17.91 | -43.85 |
| Neoplasms | 9627.33 | 3927.99 | 15891.93 | 4644.64 | 6046.11 | 12626.39 | -59.2 | -15.43 | -35.03 | -68.89 | -75.28 |
| Neurological disorders | 4260.65 | 3205.8 | 7008.13 | 2821.86 | 3362.02 | 4515.44 | -24.76 | 13.61 | -4.65 | -29 | -54.26 |
| Other non-communicable diseases | 5407.97 | 5641.05 | 4232.76 | 9073.06 | 5936.75 | 4262.3 | 4.31 | -37.83 | -4.98 | 32.35 | 33.27 |
| Sense organ diseases | 2940.03 | 1740.49 | 2898.21 | 1635.21 | 2722.44 | 3592.8 | -40.8 | 6.44 | -36.07 | -51.56 | -39.95 |
| Skin and subcutaneous diseases | 1594.72 | 1214.12 | 1853.56 | 1522.03 | 1507.09 | 1607.39 | -23.87 | -20.23 | -19.44 | -24.47 | -34.5 |
| Substance use disorders | 1235.92 | 411.8 | 3086.97 | 541.01 | 737.01 | 1203.47 | -66.68 | -23.88 | -44.12 | -65.78 | -86.66 |

### Elderly NCD Incidence Trends Over Time

Table 3. presents the change in incidence rates for both genders across the following time points: 1991, 2001, 2011, and 2021. Additionally, the table provides data on the corresponding incidence rate percentage changes per rate. Throughout the four time points, incidence rates for both genders combined remained relatively constant with only slight fluctuations over time. The incidence rate for both genders combined was 181,191.2 in 1991 reaching 181,051.2 by 2021, indicating a minimal change. The females’ and males’ incidence rates also appear to have remained relatively stable over the 4 time points. Thus, the findings suggest that the incidence of diseases has remained relatively constant in Jordan over the past three decades with only slight changes observed, according to the incidence rates.

**Table 3.** Change in incidence rates over time for individuals aged 70 years and above in Jordan (by sex).

| **Sex** | **1991** | **2001** | **2011** | **2021** | **% Change** |
| --- | --- | --- | --- | --- | --- |
| **Both** | 181191.2 | 179205.4 | 178921.4 | 181051.2 | 0.01 |
| **Female** | 186966.2 | 186091.7 | 185549.2 | 187296.5 | 0.01 |
| **Male** | 174844.8 | 172766.2 | 173040.9 | 174954.9 | 0.00 |

### Elderly NCD Mortality Trends Over Time

Figure 4. presents the death rates resulting from the most common causes of NCDs. The figure displays rankings of the various causes of death for the elderly population in Jordan from 1991 to 2021. Throughout the different time points, cardiovascular diseases appear to be the leading cause of death in Jordan. Although most of the diseases managed to remain in the same rank throughout the time period, there appears to be changes in ranks for some diseases as indicated in the figure. Rank fluctuations were shown throughout the years for both digestive diseases and chronic respiratory diseases. Digestive diseases were at Rank 5 in 1991, started dropping beginning from 2001, and finally managed to remain at Rank 6 as of 2011. On the other hand, chronic respiratory diseases were initially at Rank 6 then started increasing in 2001 to Rank 5 and remained constantly there as of 2011. The figure suggests that while some causes of death remained constant throughout the years, others have experienced significant variations in ranks possibly due to changes in public health priorities, medical advancements, and lifestyle factors affecting the elderly population in Jordan.

**Figure 4.**
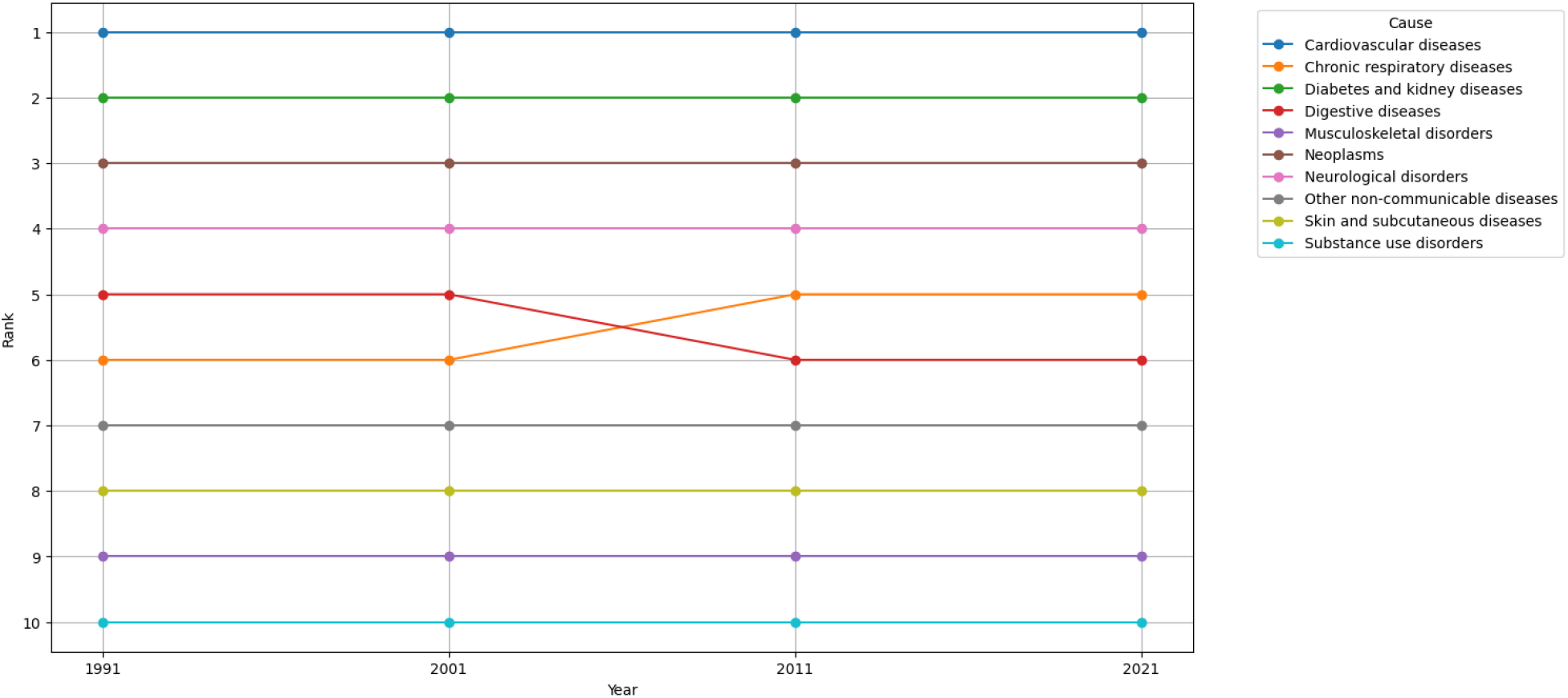
Death rate rankings of NCD causes over time.

## Discussion

With the global increase in the elderly population, it is necessary to ensure the implementation of appropriate measures to address the healthcare needs of the elderly in the aim of providing equitable public healthcare. As of 2021, the population size of the Jordanian elderly population has dramatically increased by approximately 503% relative to the year 1991.This study explores the burden of NCDs in Jordan among the elderly individuals aged 70 years and above across four time points. Additionally, this study provides a comprehensive insight and comparison of Jordan’s state of NCDs burden, across all ages, relative to the global level and income group classifications by the World Bank.

In 2021, the average burden of NCDs across all ages in Jordan was found to be 43113.17 which is 34.34% lower than the global DALYs rate per 100,000. When comparing Jordan to its similar income level countries (lower-middle income), as classified by the World Bank, Jordan appears to have a 28.35% lower average burden of NCDs. However, there are still several NCDs that need to be recognized as a public health priority given that the burden of diabetes and kidney diseases and mental disorders relative to the global level is 2.5% and 17.5%, respectively. Additionally, diabetes and kidney diseases and mental disorders in Jordan have a higher burden when compared to low-income countries of about 50% and 27%, respectively. Musculoskeletal disorders, neurological disorders, and sense organ diseases are also shown to have a higher burden in comparison to the low-income countries by 65.6 %, 13.6, and 6.4%, respectively.

Among the Jordanian elderly population in 2021, the five most current causes of NCDs burden for both genders include cardiovascular diseases, marking 39% of the total number of DALYs, diabetes and kidney diseases, neoplasms, musculoskeletal disorders, and neurological disorders.

Although the absolute numbers of DALYs for NCDs have shown a notable increase over time, from 62,840.50 in 1991 to 262,555.37 in 2021, there appears to be a decrease in the total percentage change of DALYs rate per 100,000 population of around 0.32%. This indicates that the burden of NCDs relative to the population has slightly decreased over the specified time period. However, the rate seems to increase for diabetes and kidney diseases by 0.18%, musculoskeletal disorders by 0.06%, and skin and subcutaneous diseases by 0.01% (more significantly by 0.06% for males).

The increase in the DALYs rate shown for diabetes and kidney diseases might be related to dietary habits, socio-economic factors, and accumulated exposure to the related risk factors. In addition, the increased number of kidney diseases may be due to the physiological norm of decreasing kidney function in older adults. The absolute number of DALYs for mental disorders also seemed to increase, especially in the last decade, which may reflect the positive shift in the culture’s social attitude towards seeking psychological help, indicating that mental disorders may have been underdiagnosed in the past years. The significantly increasing DALYs percentage of change for these several NCDs highlights the importance of considering them as significant health issues and prioritizing interventions in the aim of controlling and mitigating the ongoing increase in the NCDs burden over time among the Jordanian population .

The incidence rate has remained relatively constant indicating that the incidence of new cases of NCDs in Jordan among the elderly population has remained stable over the observed years of 1991 to 2021. This reflects the possibility of several factors that may have contributed including the likelihood that no significant changes in risk factors or preventive measures for NCDs have occurred during this time period. Also, early detection and management of NCDs through advancements in healthcare interventions may have had an impact on and stabilizing the incidence rate. However, it is necessary to obtain a more comprehensive understanding of the underlying reasons through conducting further research in aim to comprehend the implications of the implemented interventions on the healthcare outcomes.

Several NCDs are known to increase death rates. The results of this study show that among the Jordanian elderly population, cardiovascular diseases remained the major cause of death in comparison to other NCDs over the years. This indicates that cardiovascular diseases have consistently been a major concern among the elderly population in Jordan in line with (15). Therefore, it is essential to implement interventions to identify the factors contributing to cardiovascular diseases in aim to reduce their burden among the Jordanian population (15). While some causes of death remained constant throughout the years, digestive diseases, and chronic respiratory diseases have experienced significant variations in ranks overtime. This is potentially due to changes in healthcare policies and interventions, shifts in healthcare priorities, advancements in healthcare, and lifestyle changes including diet, physical activity, and smoking among the elderly (16). Thus, it is necessary for the healthcare sector in Jordan to implement strategies that address these issues and adjust the public health efforts accordingly in aim to decrease mortality rates among this population.

## Conclusion

The findings indicate a considerable increase in the burden of NCDs in Jordan among the elderly population making it necessary to implement comprehensive healthcare programs prioritizing chronic illness prevention, early identification, and effective management. Jordan’s healthcare system must adapt to these demographic shifts, especially due to the increasing aging population, to reduce the NCDs impact on both individuals and the economy. Future research should focus on addressing NCDs in Jordan and identifying the suitable innovative strategies through longitudinal studies in aim to track the NCD prevalence. Also, it is necessary to explore the different interventions that may be implemented to promote healthy behaviors and improve the access of the elderly to healthcare services. This can provide valuable insights for policymakers and healthcare providers in the aim of potentially enhancing the healthcare outcomes and the quality of life for Jordan’s aging population, in addition to decreasing the economic burden associated with NCDs.

## Data Availability

All data produced are available online at: https://vizhub.healthdata.org/gbd-results/

## Appendix

Population changes in Jordan over time by age groups of 70+.

| Age Group | 1991 | 2001 | 2011 | 2021 |
| --- | --- | --- | --- | --- |
| 70-74 | 24059.62508 | 39740.30266 | 83211.73647 | 141631.022 |
| 75-79 | 13446.62394 | 22185.56494 | 40246.36318 | 87764.64003 |
| 80-84 | 7774.55561 | 10862.44608 | 19210.29216 | 52954.43395 |
| 85-89 | 4136.396993 | 4420.060222 | 8824.73148 | 17566.29842 |
| 90-94 | 1241.047115 | 1455.333246 | 2701.944512 | 5410.977394 |
| 95+ | 345.1620109 | 604.0084176 | 880.0507324 | 1780.544526 |
| Total | 51003.41075 | 79267.7156 | 155075.1185 | 307107.9163 |

Source: GBD (9)

